# Short and long-term outcomes of outborn vs. inborn infants <32 weeks’ in Western Australia: A cohort study of infants born between 2005 and 2018

**DOI:** 10.1101/2022.08.13.22278651

**Authors:** J Davis, CE Seeber, E Nathan, T Strunk, A Gill, M Sharp

**Author notes:** Corresponding author: Dr Jonathan Davis, Newborn Emergency Transport Service of Western Australia, c/o Perth Children’s Hospital, 15 Hospital Avenue, Nedlands, Perth, Western Australia. Dr Seeber and Davis contributed equally to the production of the manuscript. Ethicsl review Ethical opinion was sought from the Child and Adolescent Health Service of Western Australia Institutional Review Board. Ethical approval was formally waived. Institutional approval granted as a quality improvement project by the Child and Adolescent Neonatology Quality improvement group. The institutional number given was 33045. Prior to submission for publication further approval from the review board was sought and granted by the executive director of medical services.

## Abstract

**Objective:** To compare mortality and morbidity of inborn vs outborn very preterm infants <32 weeks’ in Western Australia between 2005 and 2018.

**Design:** Retrospective cohort study

**Patients:** Infants <32 weeks’ born in Western Australia

**Main outcome measures:** Mortality was assessed as death before discharge home from the tertiary NICU. Clinically significant short-term morbidities included combined brain injury (intracranial haemorrhage (ICH) Grade ≥ 3 and cystic periventricular leukomalacia (cPVL)) and other important major neonatal outcomes. Standardised developmental assessments up to 5 years of age were evaluated where available. We performed multivariable logistic regression analysis of outborn status on outcomes, controlling for gestational age, birthweight z-score, sex and multiple birth

**Results:** A total of 4974 infants were born in WA between 22 - 32 weeks’ gestation between 2005 – 2018 of which 4237 (89.6%) inborn and 443 (10.4%) outborn were compared. Overall mortality to discharge was higher in outborn infants (20.5% (91/443) vs. 7.4% (314/4237); aOR 2.44, 95% CI 1.60-3.70, p<0.001). Outborn infants had higher rates of combined brain injury than those inborn (10.7% (41/384) vs. 6.0% (246/4115); adjusted OR 1.98, 95% CI 1.37 – 2.86), p<0.001). No difference in long-term neurodevelopmental measures was detected, however, long-term follow-up data were available for only 65% of outborn and 79% of inborn infants.

**Conclusions:** Outborn preterm infants <32 weeks in WA have increased odds of mortality, and combined brain injury than those inborn. Long-term outcome results is likely to be affected by incomplete follow-up data.

## Background

Preterm infants are at high risk of complications and death. Centralisation of perinatal care in tertiary hospitals reduces their mortality and morbidity (1-3). Infants born outside a tertiary centre and who require transport after birth have an increased risk of intracranial haemorrhage (ICH), necrotising enterocolitis, spontaneous intestinal perforation and pulmonary haemorrhage (4-9). In some populations, this has translated into long-term neurodevelopmental impairment (7, 10, 11).

Western Australia (WA) has an area of ∼2.6 million Km^2^ and population of 2.8 million people. King Edward Memorial Hospital (KEMH) is the largest perinatal centre for very preterm infants (<32 weeks’) in WA and is based in Perth, the states most populous city. Women at risk of preterm delivery in WA are transferred to KEMH for management. Outborn preterm infants are transported by the Newborn Emergency Transport Service of WA (NETS WA) by road or fixed wing aircraft to KEMH (maximum distance of travel ∼2200 km) for ongoing care. The aim of this study was to compare the short- and long-term outcomes of in- and outborn infants, 22^+0^ - 31^+6^ over a 14-year period (2005 – 2018) in WA.

## Methods

De-identified data were obtained from hospital perinatal databases and the WA midwives notification system (MNS) for patients born between 1^st^ January 2005 and 31^st^ December 2018 at 22^+0^ - 31^+6^ weeks in WA. Data from the MNS included all liveborn infants not transferred to KEMH in the study period and comprised place of birth, age (as completed weeks of gestation) and limited resuscitation information. Infants born in KEMH were referred to as ‘inborn’ and babies born anywhere outside this hospital ‘outborn’.

### Exclusion criteria

Infants were excluded from any analysis if they had lethal congenital abnormalities (consistent with similar population studies (12)) or were transferred from an institution that provided initial care but could not provide required specialist treatment such as cardiology or paediatric surgery, i.e. infants who had initial management in another centre and subsequently transferred for higher care (surgical or cardiac) input. Two other centres from 2015 onwards could care for infants born between 30 – 32 weeks. Data were not available for these infants. For long-term analysis infants were excluded if they did not meet neonatal unit follow up criteria (described below), had a non-lethal congenital anomaly or died before follow-up.

### Definitions

Outborn infants were born ‘in hospital’ if they were delivered in a medical facility with any healthcare professional readily available. Hospitals were divided into those with neonatal facilities (equipment for immediate care) and those without. A complete course of antenatal steroids was defined as the administration of >1 dose >24 hours before birth. The Australian Bureau of Statistics’ Socioeconomic Indexes for Areas (SEIFA) Index of Relative Socioeconomic Advantage and Disadvantage (IRSAD) corresponding to each mother’s address at birth was determined, to serve as an indicator of socioeconomic status. Birthweight z-scores were calculated based on national percentiles (13).

### Short-term outcomes

Overall mortality was described prior to discharge for both groups. Mortality was also compared as, death prior to transfer to a tertiary NICU for outborn infants or death prior to admission to the NICU for inborn infants in a tertiary centre. Major short-term morbidities were defined as follows: Combined brain injury was intracerebral haemorrhages (ICH) ≥ grade 3 (Papile classification) and/or cystic periventricular leukomalacia (cPVL) on ultrasound (performed and reported by the paediatric medical imaging department); Neonatal chronic lung disease (bronchopulmonary dysplasia, BPD) was a requirement for supplemental oxygen or respiratory support at 36 weeks corrected age or home oxygen on discharge, sepsis (early or late) based on positive blood culture; necrotising enterocolitis > stage II on modified Bell’s criteria, and retinopathy of prematurity (ROP) requiring laser or intraocular injection treatment (as determined by a paediatric ophthalmologist).

### Long-term outcomes

Long-term follow-up is routinely offered to infants born <28 weeks or <1000g was up to 5 years age, and 2 years corrected (CGA) for those born <30 weeks or <1250g. Medical and developmental outcome data for births between 2007 – 2016 were included. Bayley Scales of Infant and Toddler Development, 2^nd^ and 3^rd^ Ed was administered at 2 years CGA (14, 15), Griffiths Mental Developmental Scale (GMDS-Extended Revised/ER 2-8 years) at 3 years CGA (16) and Wechsler Preschool and Primary Scale of Intelligence, 3^rd^ Ed (WPPSI-III) (17) full-scale IQ (FSIQ) at age 5 years.

Severe disability was defined as cerebral palsy ≥GMFCS level 3, significant visual impairment (vision 6/60 or worse in both eyes), significant hearing impairment (requires hearing aids or cochlear implants bilaterally), 2-year-old Bayley cognitive composite <70, 3-year-old Griffiths developmental quotient <70 or 5-year-old WPPSI FSIQ <70. The latest available assessment was used to identify disability (18, 19).

### Analysis

Univariate comparisons between in- and outborn infants, within outborn infants, between metro and non-metro births and hospital type (hospital with nursery, hospital without nursery and not born in hospital) and baseline comparison of those with and without developmental assessment were made using Mann-Whitney tests for continuous outcomes and Chi-square or Fisher exact tests for categorical outcomes. The change in rate of outborn birth was compared between two epochs 2005 - 2014 and 2011 - 2018 by Mann-Whitney test. The effect of inborn vs outborn status was analysed in unadjusted and adjusted logistic regression modelling while accounting for potential confounders and covariates known to influence neonatal outcomes in preterm infants including multiple birth, gestational age, birthweight z score and sex. Receipt of antenatal steroids was not adjusted for as this is confounded by outborn vs. inborn birth. Maternal age was assessed in all models as there was a baseline imbalance. Model adjustments for neurodevelopmental outcomes included covariates listed above as well as a measure of maternal socio-economic status (lowest SEIFA quintile) and birth year. For outborn infants, the effects of time to arrival to tertiary hospital was assessed. Unadjusted (OR) and adjusted odds ratios (aOR) were reported along with their 95% confidence intervals (CI) unadjusted odds are presented when other discriminating data were not available. SPSS version 25 statistical software was used for data analysis (Armonk, NY, IBM Corp).

### Governance approval

Permissions were provided through local Women’s and Newborns Health Service governance structures (WA Health; Governance, Evidence, Knowledge, Outcomes #33045).

## Results

A total of 4974 infants were born in WA between 22-32 weeks’ gestation between 2005 – 2018; of these, 4721 were screened for analysis and were admitted to the only tertiary unit in WA during the study period, 253 infants were admitted to other appropriate institutions. Forty-one infants were excluded, a flow diagram of patients included in short and long-term analysis is presented in figure 1. There were 4237 infants inborn and 443 outborn, 85 inborn infants died before being admitted to NICU (2%) and 56 outborn infants died prior to transfer (12.6%). The analysis of short-term outcomes therefore included 4152 inborn and 387 outborn. Long-term follow-up data were available for analysis in 1370/1734 (79%) eligible surviving inborn infants and 87/133 (65%) outborn.

**Figure 1.**
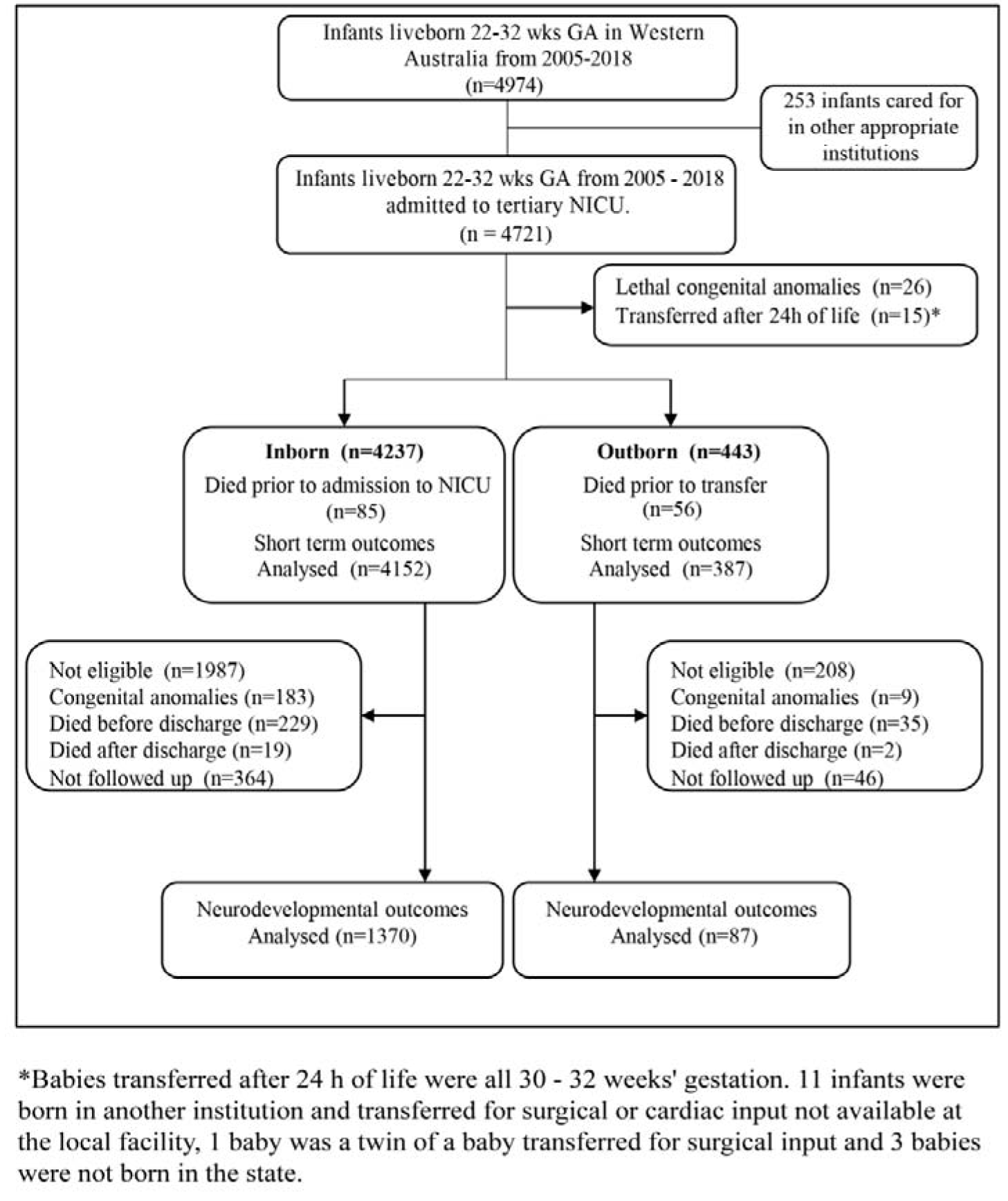
Flow diagram of preterm neonates < 32 weeks’ in Western Australia between 2005 – 2018; inclusion and exclusion criteria, breakdown of infants who died prior to admission (inborn) or transfer (outborn) and those lost to follow up.

The number of very preterm deliveries who survived to be transferred (as a proportion of all live born admissions <32 weeks’) increased from 7.4% (244/3288) from the epoch 2005 to 2014 to 11.4% (143/1251) between 2015 to 2018 (p<0.001). Outborn infants were more likely to be singleton, greater birthweight and born to younger mothers than those inborn. The mothers of outborn infants received fewer complete courses of antenatal steroid and were less likely to be delivered by caesarean section. Perinatal demographics are presented in table 1.

**Table 1.** Maternal, neonatal, labour and birth characteristics of inborn and outborn preterm neonates < 32 weeks’ in Western Australia between 2005 – 2018 admitted to KEMH NICU.

|  | **Inborn**<br>**N=4152** | **Outborn**<br>**N=387** | **p-value** |
| --- | --- | --- | --- |
| Maternal age (y); Median (IQR;R) | 30 (26-34; 14-49) | 28 (22-33; 13-46) | <0.001 |
| Gestational age (w); Median (IQR:R)<br>(Total) | 29 (27-30; 22-31) | 29 (27-30; 22-31) | 0.78 |
| <26 | 650 (15.7%) | 50 (12.9%) |  |
| 26-27 | 712 (17.1%) | 62 (16.0%) | 0.25 |
| 28-29 | 1030 (24.8%) | 111 (28.7%) |  |
| 30-31 | 1760 (42.4%) | 164 (42.4%) |  |
| Birth weight (g); Median (IQR) | 1210 (890-1515) | 1295 (1045-1610) | <0.001 |
| Birth weight z-score; Median (IQR) | 0.06 (-0.60-0.66) | 0.43 (-0.15-0.86) | <0.001 |
| Male | 2295 (55.3%) | 218 (56.3%) | 0.69 |
| Multiple birth | 1112 (26.8%) | 66 (17.1%) | <0.001 |
| Congenital anomaly | 183 (4.4%) | 9 (2.3%) | 0.05 |
| SEIFA (Lowest 2 quintiles) | 785 (19.0%) (n=4130) | 83 (21.7%) (n=383) | 0.21 |
| Antenatal steroids* |  |  | <0.001 |
| None | 160 (3.9%) | 193 (51.6%) |  |
| Incomplete | 1045 (25.2%) | 125 (33.4%) |  |
| Complete | 2943 (70.9%) | 56 (15.0%) |  |
| Mode of delivery* |  |  | <0.001 |
| Spontaneous Vaginal Delivery | 1521 (37.9%) | 222 (59.7%) |  |
| Instrumental Vaginal Delivery | 131 (3.2%) | 14 (3.6%) |  |
| Caesarean section in labour | 861 (20.8%) | 76 (19.7%) |  |
| Caesarean section not in labour | 1636 (39.4%) | 74 (19.2%) |  |

Comparison of continuous variables made by Mann Whitney U test and Chi Square for categorical.
\*Antenatal steroids unknown for 4 inborn and 13 outborn cases and mode of delivery unknown for 3 inborn and 1 outborn case.

### Mortality and short-term outcomes

The overall mortality for outborns infants was 20.5% (91/443) and 7.4% (314/4237) for inborn (unadjusted OR 3.23, 95% CI 2.50 - 4.18, p<0.001). Death *prior to transfer* to a specialist centre was 12.6% (56/443) among outborn infants and 2.0% (85/4237) for inborn infants prior to admission to NICU (unadjusted OR 7.07, 95% CI 4.97 - 10.06, p<0.001). Excluding 22-week infants, mortality for outborn infants prior to specialist centre transfer was 8.5% (36/422) and 0.8% (32/4178) for inborn infants prior to admission to NICU (unadjusted OR 12.08, 95% CI 7.42 - 19.67, p<0.001). Of those who survived to NICU, death occurred in 35/387 (9.0%) outborn and 229/4152 (5.5%) inborn infants (adjusted OR 2.44, 95% CI 1.60-3.70, p<0.001). More outborn infants had combined brain injury compared to those inborn (adjusted OR 1.98 (95% CI 1.37-2.86); p<0.001,Table 2). Mortality and short-term outcomes adjusted analysis stratified by gestation are presented in supplemental table 1. Combined brain injury and ICH ≥3 was greatest in infants with lower gestational age at birth (table 3.) A figure representing death by gestation in inborn/outborn infants who died prior to admission to NICU (inborn) or who died prior to transfer to a tertiary centre (outborn) is included in the supplement (Supplement Figure 1).

**Table 2.** Short-term outcomes as proportions of outborn and inborn infants born < 32 weeks’ in Western Australia between 2005 – 2018 admitted to KEMH NICU. Unadjusted and adjusted odds ratios (OR) and 95% confidence intervals (CI) are reported.

|  | **Inborn**<br>**n=4152** | **Outborn**<br>**n=387†** | **Unadjusted**<br>**OR (95% CI)** | **Adjusted**<br>**OR (95% CI)** | **p-value** |
| --- | --- | --- | --- | --- | --- |
| Combined brain injury<sup>#</sup> | 246/4115<br>(6.0%) | 41/384<br>(10.7%) | 1.88 (1.33-2.66) | 1.98 (1.37-2.86) | <0.001 |
| IVH grade 3/4* | 204/4119<br>(5.0%) | 31/384<br>(8.1%) | 1.69 (1.14-2.50) | 1.82 (1.20-2.76) | 0.005 |
| cPVL* | 62/4119<br>(1.5%) | 12/385<br>(3.1%) | 2.11 (1.12-3.94) | 2.00 (1.06-3.79) | 0.032 |
| BPD<sup>##</sup> | 1029/3953<br>(26.0%) | 64/358<br>(17.9%) | 0.62 (0.47-0.82) | 0.81 (0.56-1.16) | 0.250 |
| ROP requiring treatment | 121 (2.9%) | 8 (2.1%) | 0.70 (0.34-1.45) | 0.95 (0.44-2.05) | 0.891 |
| Positive blood culture | 626 (15.1%) | 56 (14.5%) | 0.95 (0.71-1.28) | 1.07 (0.78-1.46) | 0.683 |
| NEC stage 2/3 | 107 (2.6%) | 7 (1.8%) | 0.70 (0.32-1.51) | 0.88 (0.40-1.91) | 0.738 |

Short term outcomes are adjusted for multiple birth, GA, birthweight z-score and sex.
†Analysis excluded babies who died before arriving to hospital. \*Recorded for infants who had ultrasound scans. #Combined brain injury includes PVL and/or IVH grade 3/4. ##Of infants who survived to 36 weeks corrected gestational

**Table 3.**
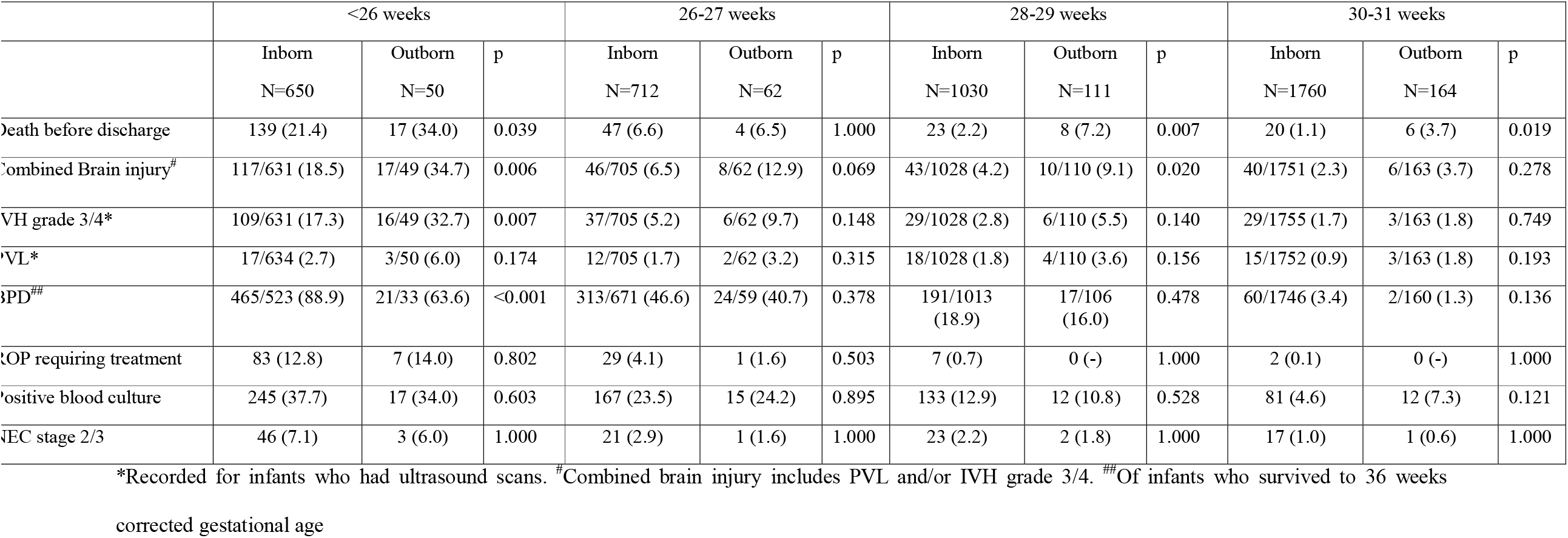
Mortality, short-term outcomes (N %) of in- and outborn infants admitted to NICU < 32 weeks’ by gestation in WA (2005 – 2018) stratified by gestational age group. Statistically significant results are in bold.

### Outborn infants

The majority of surviving outborn infants (331/387 (85.5%)) were born in a hospital with a neonatal unit, while only 17/387 (4.4%) were born in a hospital without a neonatal unit and 38/387 (9.8%) out of hospital. Of these, 17 were born en route to a medical facility, 18 were born at home, two born in communities without medical facilities and data were missing on one patient. In the population of infants who died before being transferred 51/56 (91.1%) were born in a hospital with neonatal facilities and 5/56 (8.9%) born were born in a hospital without. The median (IQR) time to arrival from birth to a tertiary perinatal centre was longer for infants born outside the Perth metropolitan area (8.5 h, 6.9-10.9) compared to those born in it (3.6 h, 3.0-4.5; p<0.001). cPVL was uncommon in the total population (74/4504 (1.6%)). The median (IQR) travel time for infants who developed cPVL was 7.1 hrs (IQR: 4.7-13.3; Range: 4.0-17.0) compared to 4.6 hrs (IQR: 3.33-7.58; Range: 0.3-56.7) in infants who did not develop cPVL (aOR 1.1, 95%CI 1.0-1.2) p=0.059).

### Long-term follow up and outcome

To estimate neurodevelopmental impairment the 2-year Bayley assessment was used in 65.1% (n=949), 3-year Griffith’s assessment in 0.3% (n=5) and 5-year WPPSI assessment in 33.8% (n=492). In the remaining 11infants (0.8%) cerebral palsy, vision or hearing assessments were used alone. Infants not followed up were more likely to be outborn to younger mothers, have lower socioeconomic status, higher gestational age and birthweight, and born by spontaneous vaginal delivery. Basic demographics of children with and without developmental assessment are shown in supplement table 2. Despite apparent trends in disability, cerebral palsy and hearing impairment there was no difference in the adjusted odds ratio (table 4).

**Table 4.** Long-term outcomes as number and proportions of outborn and inborn infants born < 32 weeks’ in Western Australia between 2005 – 2018 admitted to KEMH NICU (n=1457). Unadjusted and adjusted odds ratios (OR) and 95% confidence intervals (CI) are reported.

|  | Inborn<br>N=1370 | Outborn<br>N=87 | Unadjusted<br>OR (95% CI) | Adjusted<br>OR (95% CI) | p-value |
| --- | --- | --- | --- | --- | --- |
| Disability | 74<br>(5.4%) | 5<br>(5.7%) | 1.12<br>(0.44-2.85) | 1.08<br>(0.42-2.80) | 0.88 |
| Cognitive<br>delay (<70) | 39/1360<br>(2.9%) | 2/86<br>(2.3%) | 0.84<br>(0.20-3.55) | 0.85<br>(0.20-3.70) | 0.83 |
| Cognitive<br>score <85<br>(mild delay) | 151/1360<br>(11.1%) | 7/86<br>(8.1%) | 0.63<br>(0.27-1.46) | 0.85<br>(0.35-2.05) | 0.71 |
| Cerebral<br>palsy | 68<br>(5.0%) | 7<br>(8.0%) | 1.76<br>(0.78-3.96) | 1.74<br>(0.76-4.00) | 0.19 |
| Hearing<br>impairment | 8<br>(0.6%) | 1<br>(1.1%) | 2.01<br>(0.25-16.27) | 2.16<br>(0.25-18.95) | 0.49 |
| Vision<br>impairment | 4 (0.3%) | 0 (-) | Not tested | Not tested |  |

Long term outcomes are adjusted for multiple birth, GA, birthweight z-score, sex, low SEIFA (lowest 2 quintiles) and year of birth. Disability is defined as cerebral palsy ≥ GMFCS level 3, significant visual impairment, significant hearing impairment, 2-year-old CGA Bayley cognitive composite score, 3year old CGA Griffiths DQ or 5-year-old WPPSI FSIQ <70. Cognitive delay is defined as Bayley cognitive composite score, Griffiths DQ or WPPSI FSIQ <70. The most recent assessment was used to identify disability or delay.

## Discussion

In this population of over 4,000 very preterm infants (<32 weeks), outborn status was significantly associated with higher mortality at all points of assessment: overall, prior to transfer and after admission to NICU than those infants born in a tertiary facility. The risk of combined brain injury in outborn infants was also substantially greater than those inborn. Long-term neurodevelopmental impairment was similar in both groups, but as data were missing in a substantial proportion conclusion cannot be made.

Our data are consistent with some but not all previous reports on outborn preterm infants. (3, 10, 12, 20, 21). Total mortality from birth to discharge in the outborn group was similar to other published cohorts (10). The increase in mortality in outborn infants persists in those who survive to admission to NICU. We have shown an increase in mortality in the older gestation infants >28 weeks not demonstrated in other studies. The inborn group may be protected by greater exposure to antenatal steroids, actively managed delivery and importantly birth in a high-volume tertiary centre

Severe brain injury is reported in outborn infants and is consistent with our findings (10, 12, 21-23). In infants who require transport for specialist care after birth the underlying causes of intracranial pathology are uncertain. ICH in transported preterm infants has not been associated with vehicle type or expertise of the resuscitation team. (24) Severe brain injury (≥grade 3 ICH and parenchymal echogenicity including haemorrhagic and/or ischaemic lesions) in a transported group of Canadian infants was associated with condition at birth and immediate postnatal management requirements rather than transport (25).

Detailed data on cause of death prior to transport were unavailable in this study. Although they were born alive it is not clear if they were actively managed locally or provided with comfort care. We have included infants from 22 weeks in this study as 7 infants at 22 weeks were admitted to NICU and followed up during the time period. The majority of infants who died around the time of birth (whether inborn or outborn) were 22 weeks. Once the 22 weeks infants were excluded the odds of death were even higher for outborn infants (unadjusted OR 12.08, 95% CI 7.42 - 19.67, p<0.001). Active resuscitation of infants born at 22 weeks of gestation is a matter of great debate (26) and the limit of viability has changed during the time period of the study (27). In 2019 the British Association of Perinatal Medicine suggested that active intervention at 22 completed weeks’ gestation should be considered (28). Decisions to transfer these babies at the extreme of viability should be made with a clear understanding of the additional increased risk of mortality faced by outborn extremely preterm infants compared to those inborn.

The strengths of our study include we present a record of infants who died prior to transfer which has been missing from other comparable populations, the large number of infants reported in the study and that follow up outcomes were available. We have included important details on short term brain injury and have detailed long-term data with a good overall developmental follow up rate of 78%.

A limitation of our study is that long-term-follow-up data was missing 46/133 outborn infants (35%). In two large national cohorts, greater odds of death, neurodevelopmental and cognitive impairment have been detected in outborn infants (10, 29) with 31 - 53% follow up loss. A regional Australian study of in- and outborn infants (loss to follow-up of 43%) born <29 weeks’ between 1998 – 2004, reported no difference in neurodevelopmental outcome (21). Poor healthcare engagement, travel distance and socioeconomic factors may contribute to outborn delivery as well as poor follow up. Reasons for non-attendance to neonatal follow up are complex and conflicting. Greater distance from follow-up clinic has been identified for non-attendance (30) but increased severity of ICH is a predictor of regular attendance (31). Neonatal follow up clinics report poor compliance with variable baseline rates (30, 32-35). Infants lost to follow-up may be more likely to have disability (36, 37). We recognise that the data presented is in incomplete but comparable with other large cohorts. Improving rates of follow-up is challenging and requires substantial resources, but essential to adequately interpret long-term

The definition of neurodevelopmental impairment used does not capture many of the challenges that preterm infants may face. Future studies should consider a broader definition of the challenges for preterm survivors as there may be differences found with different definitions of disability. Finally, this study did not have data on the timing of membrane rupture and incidence of chorioamnionitis. Histological chorioamnionitis and funisitis increases the risk of brain injury (≥ grade 3 ICH and cPVL) but appears not to have impact on long-term neurodevelopment (38).

Reducing outborn birth in preterm infants requires the identification of deficiencies in healthcare education, infrastructure and barriers to transfer of mothers at risk of preterm delivery. Pre-emptive transfer to a tertiary centre and administration of antenatal steroids are essential in reducing outborn birth and associated mortality. Extremely preterm infants born in a non-tertiary hospital and transferred <48 hours have poor outcomes compared to those born in a tertiary setting(39) and in utero transport should be prioritised. Antenatal steroids given even a few hours before birth may be effective in reducing mortality (40), and in reducing the risk of ICH in at risk infants before delivery and those transported <72 h (23, 41). Survival is improved and morbidity reduced in infants <25 weeks who received antenatal steroids and should be considered even in babies at the limit of viability(42). Strategies at reducing preterm birth in WA are underway (43). Perinatal outreach education and preparation of non-specialist teams to actively manage preterm birth may improve perinatal care (10) and improve survival in this population.

## Data Availability

All data produced in the present study are contained in the manuscript

## Financial statement

This research received no specific grant from any funding agency in the public, commercial or not-for-profit sectors.

## Contribution statement

J Davis designed the original concept, interpreted results, drafted and revised the manuscript and approved for final submission.

C Seeber designed the original concept, the acquisition of data, interpreted results, revised the manuscript and approved for final submission

E Nathan assisted in the design of the original concept, acquisition and analysis of the data, revision of the manuscript and approval of final submission.

T Strunk assisted in designing the original concept, interpreting the result, revising the manuscript and approval for final submission.

A Gill assisted in designing the original concept, interpreting the result, revising the manuscript and approval for final submission

M Sharp designed the original concept, interpreted results, drafted and revised the manuscript and approved for final submission

## What is already known about this topic

- **Preterm infants <32 weeks’ gestation born outside a specialist centre have inconsistently been reported to have higher mortality than those inborn**.
- **Preterm infants <32 weeks’ gestation who are outborn have increased incidence of intraventricular haemorrhage**.

## What this study adds

- **Preterm infants <32 weeks’ gestation have a substantially greater mortality prior to and after transfer to a specialist centre, increased mortality is present in all gestational age group including more mature infants**
- **Combined brain injury was higher in outborn infants born < 32 weeks ‘compared to those inborn**

## How this study might affect research, practice or policy

- **Every effort should be made to centralise mothers in tertiary centres who have signs of preterm labour**
- **Policy and practice should focus on creating networks to support centralisation for all possible preterm births**

## Supplemental Figures and tables

Table 1. Unadjusted (OR) and adjusted (aOR) and 95% confidence intervals (CI) of mortality and important selected short-term outcomes in outborn with inborn neonatal outcomes in infants (a) <26, 26-27 and (b) 28-29- and 30-31 weeks’ gestation who were admitted to NICU in WA (2005 – 2018) stratified by gestational age group.

Figure 1. Comparison of infants in -and outborn infants stratified by gestation who either died prior to admission to NICU (inborn) or died prior to transfer (outborn) between 2005 - 2018 in Western Australia. At each gestation the respective % of infants represents the proportion of the total of inborn or outborn

Table 2. Baseline characteristics for eligible infants followed up compared with infants not followed up for neurodevelopmental assessments of outborn and inborn infants born < 32 weeks’ in Western Australia between 2005 – 2018

## Supplemental tables and figures

**Table 1.** Unadjusted (OR) and adjusted (aOR) and 95% confidence intervals (CI) of mortality and important selected short-term outcomes in outborn with inborn neonatal outcomes in infants <26, 26-27 (a) 28-29- and (b) 30-31 weeks’ gestation who were admitted to KEMH NICU in WA (2005 – 2018) stratified by gestational age group.

|  | <26 weeks<br>N=700 |  |  |  | 26-27 weeks<br>N=774 |  |  |  |
| --- | --- | --- | --- | --- | --- | --- | --- | --- |
|  | OR (95%CI) | p | aOR (95% CI) | p | OR (95%CI) | p | aOR (95% CI) | p |
| *Short term* |  |  |  |  |  |  |  |  |
| Died before discharge | 1.89 (1.02-3.45) | 0.042 | 2.22 (1.14-4.35) | 0.020 | 0.97 (0.34-2.78) | 0.964 | 1.96 (0.64-5.88) | 0.242 |
| Combined brain injury<sup>#</sup> | 2.33 (1.25-4.35)<br>(n=680) | 0.008 | 2.60 (1.34-5.04) | 0.005 | 2.12 (0.95-4.73)<br>(n=767) | 0.065 | 1.92 (0.83-4.42) | 0.126 |
| BPD<sup>##</sup> | 0.22 (0.10-0.47)<br>(n=556) | <0.001 | 0.27 (0.12-0.59) | 0.001 | 0.78 (0.46-1.35)<br>(n=730) | 0.379 | 1.17 (0.65-2.08) | 0.604 |

|  | 28-29 weeks |  |  |  | 30-31 weeks |  |  |  |
| --- | --- | --- | --- | --- | --- | --- | --- | --- |
|  | N=1141 |  |  |  | N=1924 |  |  |  |
|  | OR (95%CI) | p | aOR (95% CI) | p | OR (95%CI) | p | aOR (95% CI) | p |
| *Short term* |  |  |  |  |  |  |  |  |
| Died before discharge | 3.45 (1.49-7.69) | 0.004 | 3.33 (1.43-7.69) | 0.005 | 3.33 (1.30-8.33) | 0.011 | 3.13 (1.22-7.69) | 0.018 |
| Combined brain injury<sup>#</sup> | 2.29 (1.12-4.70)<br>N=1138 | 0.024 | 1.80 (0.86-3.75) | 0.120 | 1.64 (0.68-3.92)<br>N=1914 | 0.270 | 1.53 (0.63-3.70) | 0.344 |
| BPD<sup>##</sup> | 0.82 (0.48-1.41)<br>(n=1119) | 0.479 | 1.16 (0.65-2.07) | 0.626 | 0.36 (0.09-1.47)<br>(n=1906) | 0.153 | 0.41 (0.10-1.70) | 0.216 |

Short term outcomes are adjusted for multiple birth, GA, birthweight z-score and sex. <sup>#</sup> Combined brain injury includes PVL and/or IVH grade 3/4. <sup>##</sup>Of infants who survived to 36 weeks corrected gestational age.

**Figure 1.**
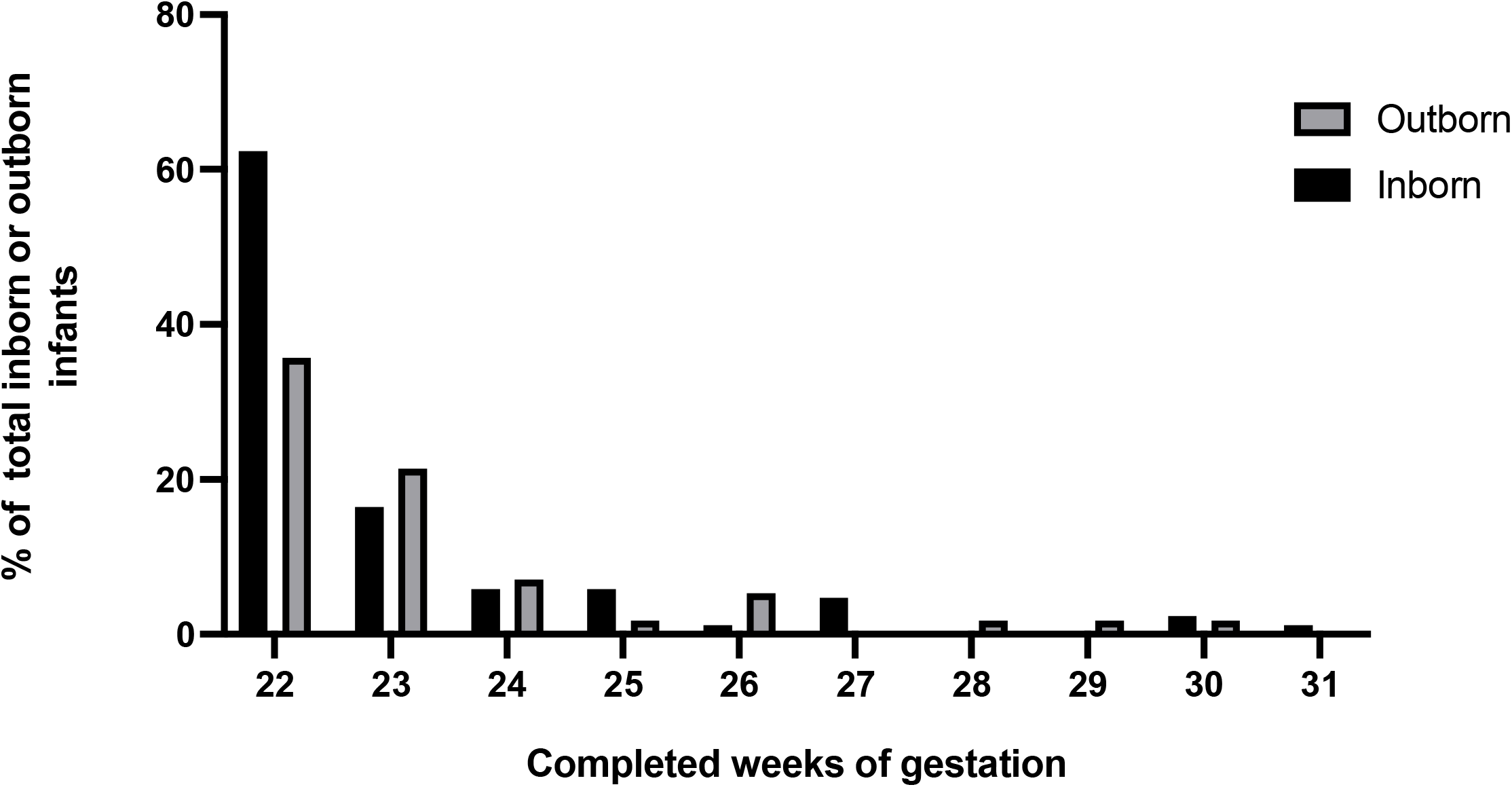
Comparison of infants in -and outborn infants stratified by gestation who either died prior to admission to NICU (inborn) or died prior to transfer (outborn) 2005 - 2018 in Western Australia. At each gestation the respective % of infants represent the proportion of the total of inborn or outborn deaths.

**Table 2.** Baseline characteristics for eligible infants followed up compared with infants not followed up for neurodevelopmental assessments of outborn and inborn babies born < 32 weeks’ in Western Australia between 2005 – 2018

|  | **Followed up**<br>**N=1457** | **Not followed up**<br>**N=410** | **p-value** |
| --- | --- | --- | --- |
| Inborn | 1370 (94.0%) | 364 (88.8%) | <0.001 |
| Maternal age, Median (IQR; R) | 30 (26-34;13-48) | 28 (23-33;16-46) | <0.001 |
| SEIFA (Lowest 2 quintiles) | 196/1445 (13.6%) | 136/409 (33.3%) | <0.001 |
| Regional area | 30 (2.1%) | 27 (6.6%) | <0.001 |
| Antenatal steroids |  |  | 0.21 |
| None | 92 (6.3%) | 35 (8.6%) |  |
| Incomplete | 366 (25.1%) | 93 (22.7%) |  |
| Complete | 999 (68.6%) | 281 (68.7%) |  |
| Multiple birth | 360 (24.7%) | 106 (25.9%) | 0.64 |
| Mode of delivery |  |  | 0.05 |
| SVD | 516 (35.5%) | 173 (42.2%) |  |
| Instrumental VD | 47 (3.2%) | 7 (1.7%) |  |
| CS in labour | 318 (21.9%) | 83 (20.2%) |  |
| CS not in labour | 574 (39.5%) | 147 (35.9%) |  |
| Gestational age (w) Median (IQR; R) | 27<br>(26-29;22-31) | 28<br>(26-29;23-31) | <0.001 |
| Birth weight (g) Median (IQR;R) | 1010<br>(810-1200; 380-1850) | 1110<br>(880-1249;420-2060) | <0.001 |
| Birth weight z-score Median (IQR;R) | -0.06<br>(-0.77-0.70; -3.08-3.64) | -0.04<br>(-0.70-0.67; -2.92-3.35) | 0.89 |
| Male | 787 (54.0%) | 223 (54.4%) | 0.89 |

## Notes

### Competing Interest Statement

The authors have declared no competing interest.

### Funding Statement

The study did not receive any funding

### Author Declarations

Ethical opinion was sought from the Child and Adolescent Health Service of Western Australia Institutional Review Board. Ethical approval was formally waived.

